# Rapid Detection of Inter-alpha Inhibitor Proteins in Neonatal Sepsis and Necrotizing Enterocolitis

**DOI:** 10.64898/2026.01.30.26345077

**Authors:** Ray H Chen, Birju A Shah, Hala Chaaban, Jessica Schuster, Richard Tucker, Andre Santoso, Joseph Qiu, Nicholas G Guerina, Yow-Pin Lim, James F Padbury

## Abstract

Inter-alpha Inhibitor Proteins (IAIP) are serine protease inhibitors and a reliable inflammatory biomarker. We have demonstrated that IAIP levels decrease during sepsis in humans and animal models. Currently, enzyme-linked immunosorbent assay (ELISA) is the standard procedure to measure IAIP levels. We developed a lateral flow immunoassay (LFIA) that allows rapid, point of care detection. We compared IAIP levels in infants with sepsis and/or necrotizing enterocolitis (NEC) by ELISA and LFIA in a multi-center, cross-sectional study. Blood samples were collected from 47 infants with sepsis, 31 sepsis case controls, 52 gestational age (GA)-matched controls and 10 infants with culture-negative sepsis (“clinical sepsis”). We also collected samples from 17 infants with NEC, 7 NEC case controls and 15 GA controls. IAIP levels at presentation of acute events and over the next 72 hours were significantly reduced in infants with sepsis, NEC and culture negative sepsis when compared to controls. IAIP levels did not differ in patients with sepsis or culture negative sepsis. IAIP levels measured by ELISA and LFIA were highly correlated (R^2^ = 0.9326) and both showed reliable detection of neonatal sepsis, NEC and culture negative sepsis. IAIP levels were 80.0% sensitive and 92.3% specific using LFIA for the detection of neonatal sepsis. For detection of NEC, IAIP levels were 84.6% sensitive and 86.7% specific.

## Introduction

Sepsis and necrotizing enterocolitis (NEC) are among the leading causes of morbidity and mortality in the newborn period^1,2^. Sepsis encompasses a systemic inflammatory response to overwhelming infection of the bloodstream. Infants with NEC present with injury of the intestinal tract and systemic inflammatory responses. Both conditions are associated with temperature instability, apnea, bradycardia, lethargy, hypotension, disseminated intravascular coagulation (DIC) and combined respiratory and metabolic acidosis^3^. In reported series, 40% to 60% of confirmed NEC cases also have concurrent sepsis^4,5,6^. The mortality rate from sepsis among very low birth weight infants is 15-20%. Moreover, the overall mortality rate of NEC among very low birth weight infants is 25-30%^7^. Early recognition and treatment of sepsis and NEC are associated with improved outcomes^7–8^.

A major limitation in the diagnosis of sepsis and NEC is the lack of a single reliable biomarker that can effectively differentiate these conditions from other neonatal illnesses. Given their rapid progression and high morbidity, there is an urgent need for accurate and timely diagnostic tools to guide clinical decision-making. This is particularly important for NEC, where early identification could help reduce unnecessary treatment, including prolonged antibiotic exposure and interruptions in enteral feeding. A more precise diagnostic tool could help balance the risks of delayed NEC diagnosis with the potential harms of overtreatment.

Inter-alpha inhibitor proteins (IAIP) are key components of the innate immune system and have been investigated as inflammatory biomarkers^9–14^. These serine protease inhibitors are produced in the liver and circulate in blood at relatively high concentrations. IAIP levels are rapidly reduced during immune system activation associated with sepsis. This is the result of suppressed hepatic synthesis, cleavage of the light chain, and renal excretion^15^. There are distinct mechanisms in the inflammatory pathways associated with sepsis that contribute to the beneficial effects of IAIP including: 1) inhibition of destructive serine proteases such as elastase and cathepsin G^16^; 2) down-regulation of pro-inflammatory cytokines such as TNF-alpha and IL-6^17–18^; and 3) blocking excess complement activation and generation of circulating C5a^19^. IAIP also bind to Danger-associated Molecular Patterns (DAMPs) such as High Mobility Group Box-1 (HMGB1)^20^, lipopolysaccharide and extracellular histones^21^.

Researchers have used an established laboratory-based enzyme-linked immunosorbent assay (ELISA) technique to measure IAIP levels, relying on a monoclonal antibody targeting the light chain^22^. However, ELISA is time-intensive, requiring trained personnel and several hours for completion. To address this limitation, we have developed a lateral flow immunoassay (LFIA) ^23^ for rapid, point-of-care IAIP measurement that provides results in less than 15 minutes. We previously demonstrated, using an ELISA procedure, that infants with sepsis and NEC have reduced IAIP levels due to associated systemic immune activation^12,24^. We also have demonstrated the high diagnostic reliability of measurement of IAIP levels in discrimination of spontaneous intestinal perforation (SIP) from NEC^25^. In this study, we aimed to confirm our prior findings and evaluate LFIA as a rapid diagnostic tool by directly comparing it to ELISA. We hypothesized that LFIA could provide accurate blood IAIP levels in these infants as well. We conducted a multi-center, prospective, cross-sectional study to assess IAIP levels in neonates with sepsis, NEC and case controls to validate LFIA’s performance in the different clinical groups.

## Methods

### Subject identification

#### Sepsis and NEC

The study included infants admitted to the NICUs at Women & Infants Hospital of Rhode Island (WIH) and the University of Oklahoma (OU) Health Sciences Center Oklahoma Children’s Hospital between August 2019 and November 2023. Following informed consent, residual blood samples were collected from infants with sepsis, NEC, case controls, and gestational age (GA) matched control infants. The Institutional Review Boards of WIH (Project ID: 11-0124) and OU (Project ID: 9393) approved the collection and use of de-identified clinical information. Residual samples were obtained at the time of an acute event, defined as the point when clinicians ordered blood cultures and other laboratory tests in response to the subject’s presenting signs/symptoms. Additional samples were collected over the following 14 days, when available. Neonatal sepsis was defined as positive blood in neonates with clinical signs of infection, requiring a full course of antibiotics (5-14 days)^26^. NEC was defined as modified Bell’s stage ≥ 2^27^. To reflect real-world clinical practice, case controls were selected based on initial clinical suspicion of sepsis or NEC, followed by subsequent exclusion through diagnostic evaluation. Infants in the case control groups underwent laboratory testing, empiric antibiotic therapy, and clinical monitoring, but were ultimately determined not to have NEC or sepsis based on negative blood cultures, normal imaging, symptom resolution, and overall clinical course. Additionally, gestational age (GA)-matched controls were recruited within ±2 weeks of birth GA from healthy, asymptomatic infants with no signs of infection or intestinal disease for comparative analysis. Infants with chromosomal or major malformations were excluded from the study. Blood samples were retrieved from the clinical lab within 72 hours of collection, centrifuged for plasma or serum separation, and stored at −80°C until analysis.

#### “Clinical sepsis”

To reflect real-world clinical practice, infants with a diagnosis of “clinical sepsis” (also referred to as “culture-negative neonatal sepsis” or “presumed sepsis”) were also enrolled in this study. These infants presented with signs of infection and whose illness severity or initial laboratory evaluations were so abnormal, the clinical providers chose to administer a full course of antibiotics despite negative blood cultures.

### IAIP Measurement

#### Enzyme-Linked Immunosorbent assay (ELISA)

IAIP were quantitatively measured by an ELISA using a human monoclonal antibody specific for the light chain of human IAIP as a capture antibody (MAb 69.26, ProThera Biologics, Providence, RI). Briefly, 50 ng of purified IgG of MAb 69.26 in phosphate-buffered saline (PBS) was immobilized on Microlon 600 High Binding 96-well microplate (Greiner BioOne, Monroe, NC) for 1 h at room temperature (RT). Serial dilutions of human plasma with known IAIP concentration in PBS were used to establish a standard curve. Fifty μl of plasma samples diluted 1:100 in PBS or serially diluted IAIPs were added to individual wells of a 96 well plate to quantify the IAIP levels. Subsequently, 50 μl of biotinylated heparin was added to each well, and the plates were incubated on a shaker for 30 min at room temperature, followed by three washes with PBS and 0.05 Tween 20 (PBS-T). Subsequently, 50 μl 1:2000 diluted biotin-conjugated Heparin was added to the well. Heparin sodium salt from porcine intestinal mucosa (Cat# SRE0027) was obtained from Sigma-Aldrich (St. Louis, MO) and biotin was conjugated to heparin by using Biotin-Hydrazide (ApexBio, Houston, TX) and a modified protocol by Frost *et al*^28^. Briefly, heparin was dissolved in 0.1 M MES buffer, pH 4.0 and biotin-hydrazide dissolved in DMSO was added to a final concentration of 1 mM. A stock solution of 1-ethyl-3-(3-dimethylamin-opropyl) carbidodiimide (EDAC) was prepared as a 100 mM stock solution in dH2O and added to the heparin–biotin solution at a final concentration of 30 mM. The free biotin and EDAC were removed using Amicon ultrafiltration device with MWCO 3 kDa (Millipore Sigma, Burlington, MA). The biotinylated heparin was aliquoted and stored in PBS containing 50% glycerol and kept in the −30°C freezer.

The bound biotinylated heparin to captured IAIP was detected by adding HRP-conjugated streptavidin (1:5000 dilution, Innova Biosciences, Cambridge, UK) for 30 min at RT. After three washes with PBS-T (PBS + 0.5% Tween 20), 100 μl Enhanced K-Blue TMB substrate (Neogen, Lexington, KY) was added, and the reaction was then stopped by adding 100 μl 1 M HCl. The absorbance at 450 nm was measured using SpectraMax reader (Molecular Devices, Freemont, CA). Each plasma sample was assayed in triplicate and the mean value was calculated. The linear range of the assay was between 50 and 1000 μg/ml and the inter and intra-assay variations were determined to be less than 5%.

#### Quantitative Lateral Flow Immunoassay (LFIA)

The established ELISA was adopted to a rapid test lateral-flow immunoassay format by using the heparin-conjugated nanogold particles. Heparin was passively absorbed to 40 nm nanogold particles (DCN Dx, Carlsbad, CA) in 5 mM NaHCO_3_ buffer containing 150 mM NaCl at pH 9.6. Following the conjugation, the nanoparticles were blocked with BSA and stored at 20 OD in dH_2_O until use. Lateral Flow Materials kit containing the complete components to assemble the lateral flow assay such as nitrocellulose membrane, conjugate, sample and absorbent pads, adhesive backing support, test strip housing was obtained from DCN Dx (Carlsbad, CA). Purified IgG of MAb 69.26 (ProThera Biologics, Providence, RI) at 1mg/mL and the purified bovine IAIP at 1 mg/mL were dispensed to the nitrocellulose membrane at the test line and control line using Biodot XYZ 3060 instrument (Irvine, CA). The membrane was previously assembled to the card base using a vacuum assembly machine Matrix 2210 (Kinematics Automation, Sonora CA). The stripped card was then dried in the oven at 37°C for 4 hours. Tape the edge of the card so the card strips will remain flat inside the oven. Following the drying, the wick pad and the conjugate/sample pad were assembled under vacuum using the Matrix 2210 instrument. The assembled card was then cut with 5 mm width using the cutting machine CM4000 (Biodot, Irvine, CA). The test strips were then inserted to the plastic housing cassettes and store in the vacuum bag and silica gel pouch at room temperature until use.

To run the test, 4 μL of the heparin-conjugated gold nanoparticles was added to 1 μL of patient’s plasma sample or the standard plasma with known IAIP concentration in a 1.5 mL tube. Additional 70 μL running buffer (PBS + 1% Tween 80) was added to the tube. Following gently mixing, all the mixed solution was transferred to the sample well of the test cassette that lied horizontally flat. After 11 minutes, the control and test lines were apparent and the cassette was placed in the tray of the iPeak reader (IUL Inc., Alexandria, VA) for quantification of the test line intensity. The optical density (OD) value displayed on the iPeak reader was then plotted against the OD of the plasma standard curve to calculate the IAIP concentration.

### Statistics

We used T-test, Chi-squared test and Fisher’s exact test for bivariate and multivariate evaluation. Statistical significance was defined as p-value < 0.05. We used R version 4.4.1 to calculate the receiver operating characteristic (ROC) curve, area under the curve (AUC), threshold, sensitivity, specificity, accuracy, positive and negative predictive values, and 95% confidence interval of the IAIP LFIA. We used SAS version 9.4 (SAS Institute, Cary, North Carolina, USA) for a multivariate regression analysis including stepwise regressions and single step models. We started by selecting significant variables that had a correlation with a p-value of 0.2 for any of the IAIP measures, with any that ended up with a p-value of 0.05 being retained. Group (sepsis, NEC and “clinical sepsis” vs controls) was forced into the models and retained whether significant or not. The other variables that were tested in all of the models were birth weight, weight at acute events, GA, gender and IVH. After the selection procedure was run, we ran single step models with the retained variables.

## Results

Forty-nine infants with culture-positive sepsis, thirty-one case controls and 52 gestational age (GA) controls were enrolled for comparison to the infants with sepsis. Seventeen infants with confirmed NEC, 7 case controls and 15 GA controls were enrolled for the comparison to the infants with NEC. Additionally, ten infants with “clinical sepsis” were enrolled. Table 1 summarizes the clinical and demographic characteristics of the study population, which were largely similar with two exceptions. As expected, infants diagnosed with sepsis and NEC had longer periods of not receiving enteral feeds. The infants with sepsis and NEC had longer periods of feeding withdrawal (2.6 vs 0.3 d, p-value = 0.02 and 12.3 vs 2.6 d respectively, p-value = 0.006). They also received a longer duration of antibiotic treatment (10.4 vs 2.3 d, p < 0.0001 and 9.8 vs 2.1 d, p = 0.001). The characteristics of infants with clinical sepsis are shown in Table 2. Infants in this group received antibiotics for an average of 6.6 days. The mean duration of cessation of feeds in this group was 3.2 days. Six of the10 infants with ‘clinical sepsis’ exhibited leukocytosis.

**Table 1.** Infant characteristics.

|  | Sepsis groups |  |  |  | NEC groups |  |  |  | “Clinical sepsis”<br>(n = 10) |
| --- | --- | --- | --- | --- | --- | --- | --- | --- | --- |
|  | Sepsis<br>(n = 49) | Case control<br>(n = 31) | GA control<br>(n = 52) | p-value | NEC<br>(n = 17) | Case control<br>(n = 7) | GA control<br>(n = 15) | p-value |  |
| Maternal age, mean (SD), y | 28.2 (5.4) | 29.4 (5.2) | 29.1 (6.6) | 0.6414 | 29.8 (5.1) | 32.1 (8.9) | 30.9 (5.7) | 0.6902 | 28.7 (7.7) |
| Term birth, No. (%) | 13 (27) | 9 (29) | 12 (23) | 0.8270 | 0 (0) | 1 (14) | 1 (7) | 0.3117 | 4 (40) |
| Gestational age, mean (SD), w | 30.8 (5.9) | 32.1 (5.5) | 32.3 (5.1) | 0.3646 | 29.1 (3.9) | 29.1 (4.2) | 31.0 (4.3) | 0.3805 | 31.1 (5.7) |
| Birth weight, mean (SD), g | 1818.6 (1243.2) | 1846.1 (1105.8) | 1864.1 (1100.4) | 0.9807 | 1306.6 (649.2) | 1428.6 (722.1) | 1623.7 (836.4) | 0.4856 | 1973.2 (1336.1) |
| Weight at acute event, mean (SD), g | 2226.2 (1387.8) | 2231.5 (1139.8) | N/A | 0.9860* | 1584.4 (662.9) | 1605.7 (668.7) | N/A | 0.9437* | 2533.7 (1577.5) |
| Small for gestational age, No. (%) | 6 (12) | 7 (23) | 7 (13) | 0.4127 | 2 (12) | 0 (0) | 3 (20) | 0.5784 | 0 (0) |
| Female, No. (%) | 20 (41) | 13 (42) | 26 (50) | 0.1755 | 7 (41) | 2 (29) | 9 (60) | 0.4093 | 6 (60) |
| Cesarean delivery, No. (%) | 27 (55) | 20 (65) | 37 (71) | 0.2441 | 13 (76) | 4 (57) | 8 (53) | 0.3467 | 7 (70) |
| ROM ≥ 18 h, PROM or PPROM, No. (%) | 9 (18) | 5 (16) | 14 (27) | 0.4210 | 5 (29) | 3 (43) | 4 (27) | 0.8095 | 0 (0) |
| Maternal diabetes, No. (%) | 5 (10) | 3 (10) | 6 (12) | 0.9608 | 1 (6) | 2 (29) | 1 (7) | 0.2828 | 0 (0) |
| Maternal GBS colonization, No. (%) | 6 (12) | 8 (26) | 12 (23) | 0.2429 | 2 (12) | 2 (29) | 2 (13) | 0.6192 | 1 (10) |
| Antenatal steroids, No. (%) | 27 (55) | 19 (61) | 28 (54) | 0.7906 | 12 (71) | 4 (57) | 8 (53) | 0.6237 | 3 (30) |
| Postnatal steroids, No. (%) | 9 (18) | 2 (6) | 5 (10) | 0.2187 | 2 (12) | 0 (0) | 3 (20) | 0.5784 | 1 (10) |
| Surgical NEC, No. (%) | N/A | N/A | N/A | N/A | 1 (6) | N/A | N/A | N/A | N/A |
| Feed withdrawal period, mean (SD), d | 2.6 (5.4) | 0.3 (0.7) | N/A | 0.0213* | 12.3 (8.4) | 2.6 (0.5) | N/A | 0.0064* | 3.2 (3.3) |
| Days on antibiotics, mean (SD), d | 10.4 (7.2) | 2.3 (0.9) | N/A | < 0.0001* | 9.8 (5.3) | 2.1 (0.4) | N/A | 0.001* | 6.6 (2.0) |

GA: gestational age; GBS: group B streptococcus; ROM: rupture of membranes; PROM: premature rupture of membranes; PPROM: preterm premature rupture of membranes
\*sepsis vs case control; \*NEC vs case control

**Table 2.** Infants with “clinical sepsis”.

| GA | PMA at acute event | Hx, manifestations, labs | Rx | Days on antibiotics |
| --- | --- | --- | --- | --- |
| 33+0 | 33+6 | neutropenia | ampicillin, gentamicin, piperacillin/tazobactam | 7 |
| 27+0 | 27+0 | severe maternal chorioamnionitis with necrotizing funisitis (all vessels involved), extreme leukocytosis | ampicillin, gentamicin | 7 |
| 27+5 | 29+2 | shock, leukocytosis | vancomycin, cefotaxime | 5 |
| 38+1 | 38+1 | shock, leukocytosis, elevated CRP | ampicillin, amikacin | 10 |
| 39+6 | 44+6 | worsening respiratory status requiring intubation, shock requiring pressors | vancomycin, ceftazidime, piperacillin/tazobactam | 5 |
| 37+2 | 37+2 | worsening respiratory status requiring intubation | ampicillin, amikacin, ceftazidime | 5 |
| 27+6 | 45+1 | leukocytosis, bandemia, elevated CRP | vancomycin, amikacin, ceftazidime | 10 |
| 24+2 | 33+2 | persistent ileus | vancomycin, amikacin | 7 |
| 31+1 | 31+1 | leukocytosis | ampicillin, amikacin | 5 |
| 25+0 | 25+0 | leukocytosis | ampicillin, amikacin | 5 |

PMA: post-menstrual age; CRP: C-reactive protein

IAIP levels at the time of acute events are shown in Figure 1. IAIP levels measured by ELISA were significantly lower in infants with sepsis and NEC compared to case controls and GA controls (*p* < 0.05). Similarly, infants with clinical sepsis had significantly lower IAIP levels than Case Controls or GA controls in the sepsis group (*p* < 0.05). Similarly, IAIP levels measured by LFIA demonstrated significantly lower values in infants with sepsis, NEC, and clinical sepsis groups compared to Case Controls or GA controls. The correlation between IAIP measurements by ELISA and LFIA was strong (R² = 0.9326, Figure 2), supporting LFIA as a reliable and rapid alternative to ELISA for IAIP quantification. Serial IAIP levels in the available samples 72 hours following acute events are shown in Figure 3. IAIP levels in NEC and clinical sepsis cases were consistently lower than in case controls at 24h, 48h, and 72h when measured by both ELISA and LFIA, though the LFIA measurement at 24 hours did not reach statistical significance.

**Figure 1.**
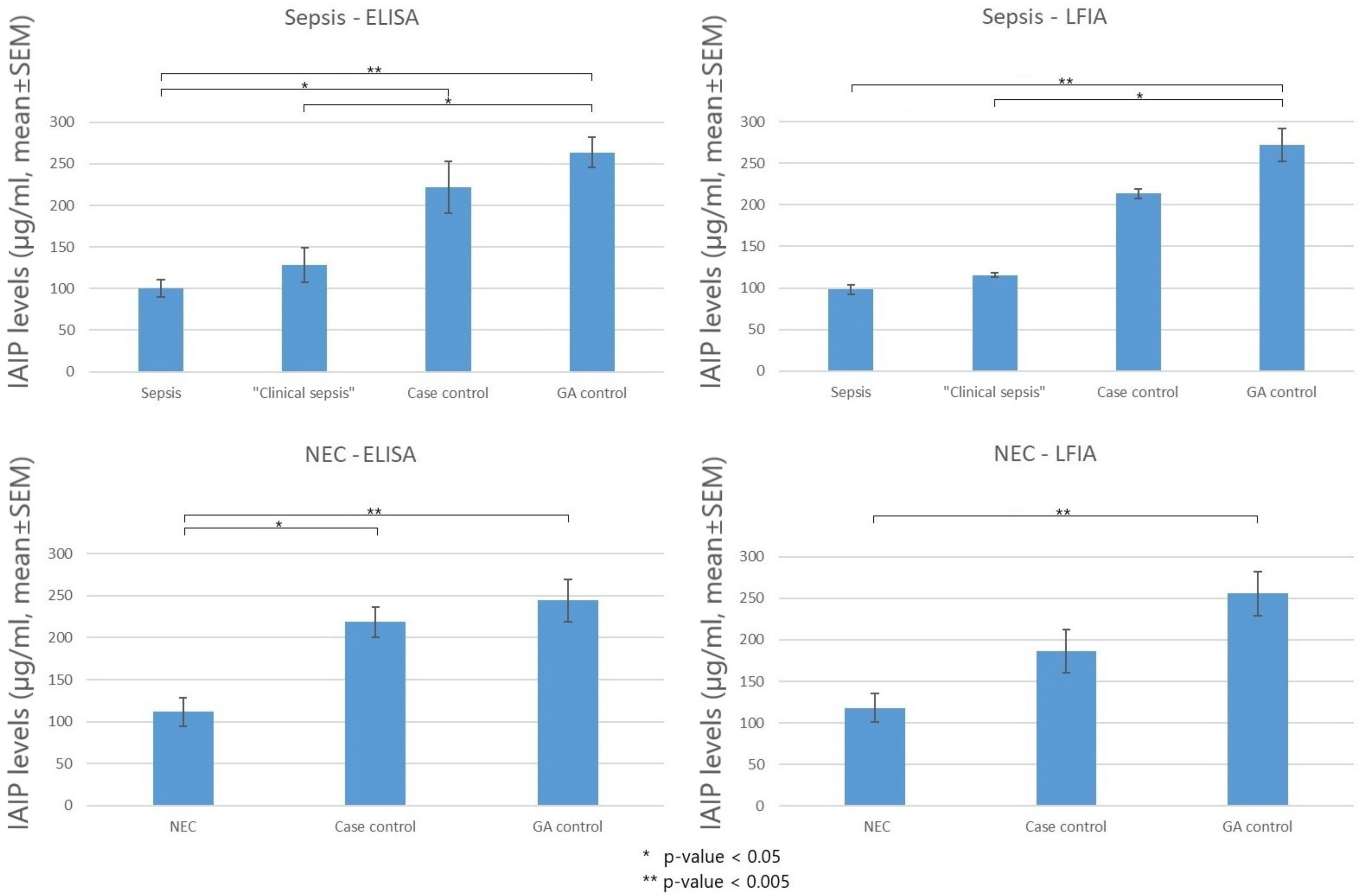
IAIP levels measured at acute events by ELISA and LFIA in infants with sepsis, clinical sepsis, NEC, case controls and GA controls. Case definitions and ELISA and LFIA assays are described in Methods. Mean ± SEM.

**Figure 2.**
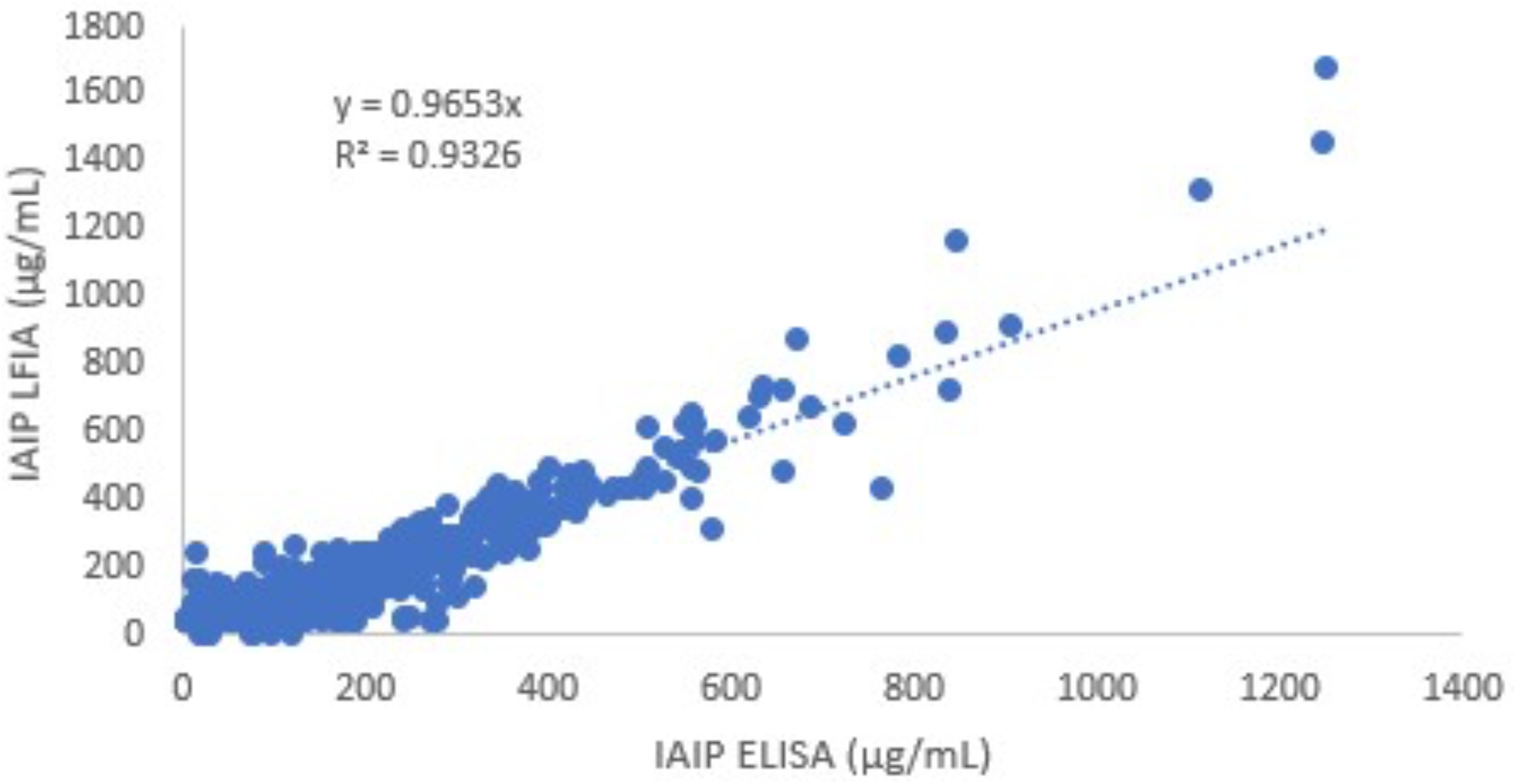
Correlation of IAIP measurements by ELISA and LFIA. R^2^ = 0.9326.

**Figure 3.**
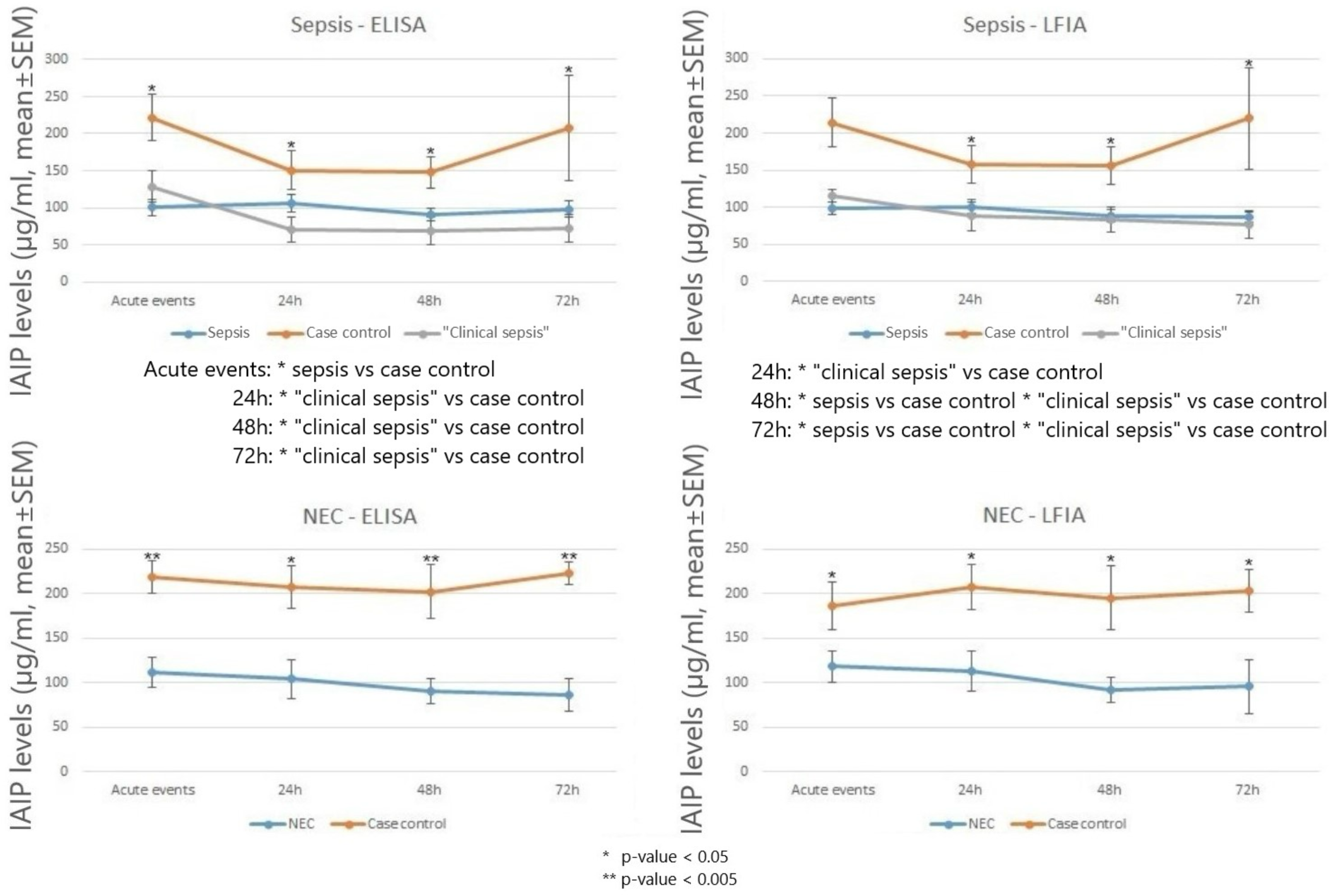
IAIP levels in the first 72 hours after acute events in infants with sepsis, clinical sepsis, NEC and case controls. Case definitions and ELISA and LFIA assays are described in Methods. Mean ± SEM.

Figure 4 shows the receiver operating characteristic (ROC) curves for the diagnosis of sepsis and NEC by ELISA and LFIA. Table 4 summarizes the main performance indicators of IAIP ELISA and LFIA. The area under the curve (AUC) for sepsis detection was 0.893 for ELISA and 0.925 for LFIA, indicating strong discriminatory power for both methods. For NEC, the AUC was 0.872 for ELISA and 0.887 for LFIA, showing similarly high accuracy. For neonatal sepsis, IAIP measured by LFIA had a sensitivity of 80.0% and a specificity of 92.3%, with a positive predictive value (PPV) of 87.5% and a negative predictive value (NPV) of 87.3%. For NEC detection, IAIP measured by LFIA had a sensitivity of 84.6% and a specificity of 86.7%, with a PPV of 84.6% and an NPV of 86.7%. These values indicate IAIP’s strong potential as a biomarker for early identification of neonatal sepsis and NEC, with LFIA providing rapid and comparable results to ELISA.

**Figure 4.**
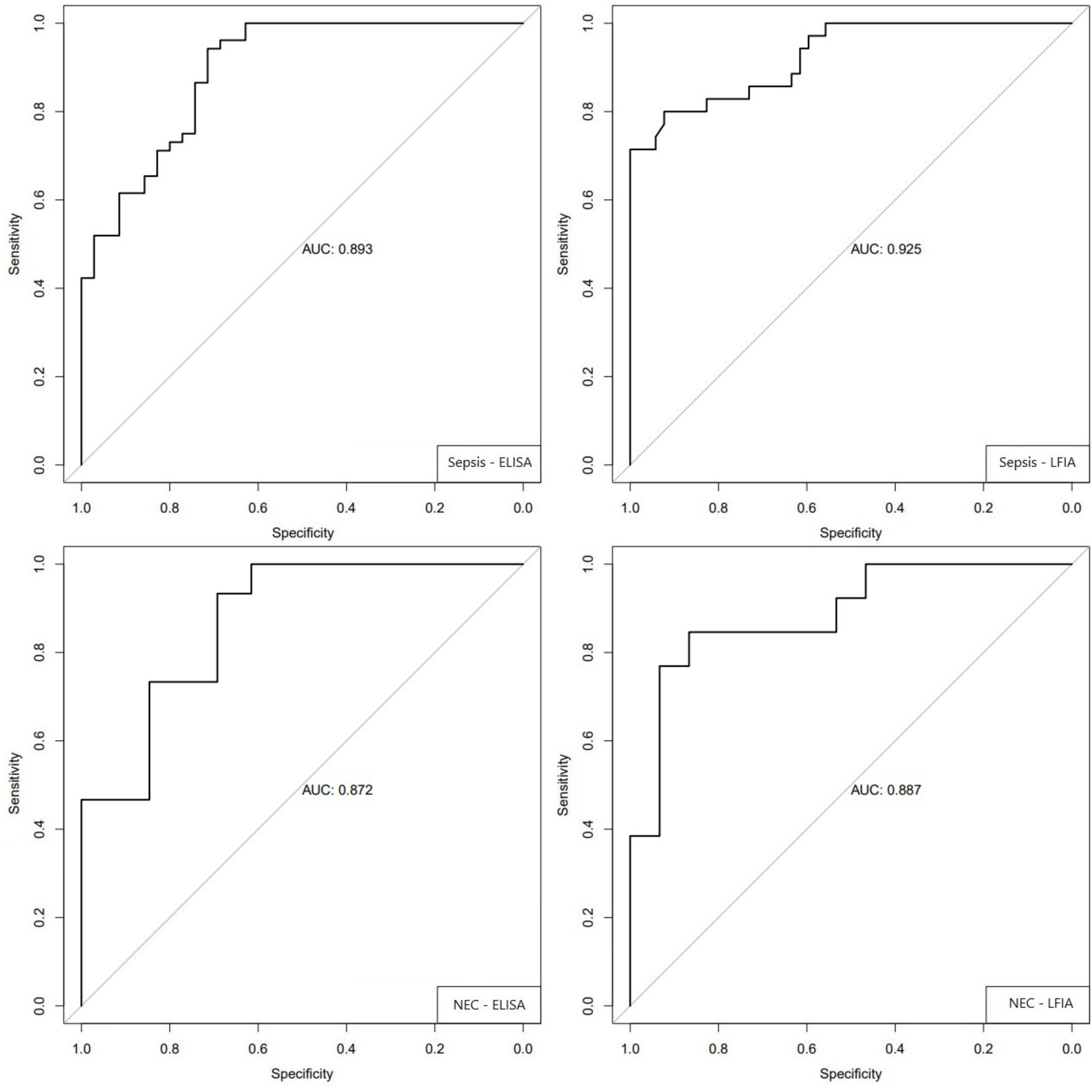
Receiver operating characteristic (ROC) curves for the diagnosis sepsis and NEC. Measurements of IAIP were made by ELISA and LIFA as shown in the inset. Area under each Reciever Operating Curve (AUC) is shown in each figure.

## Discussion

Rapid detection of blood analytes and biomarkers by LFIA has become increasingly popular^23^. This study compared of measurement of IAIP by ELISA and LFIA in infants with confirmed sepsis, confirmed NEC, “clinical sepsis” and controls. We demonstrated high diagnostic reliability for LFIA when compared to the reference ELISA results, with very high sensitivity, specificity, positive and negative predictive values. In the blood IAIP circulate in relatively high concentration as a heterotrimeric protein complex. There are two forms: Pre-alpha Inhibitor (*PaI*) and Inter-alpha Inhibitor (*IaI*). *PaI* contains one heavy chain (H3) and a light chain and has a molecular weight of 125 kDa. *IaI* is composed of 2 heavy chains (H1, H2) and a light chain and has a molecular weight of 250 kDa. The light chain, also known as bikunin, contains two Kunitz protease inhibitor domains and functions as a serine protease inhibitor^17^. The heavy chains play a role in tissue homeostasis^18^. Bikunin is bound to the heavy chains by glycosaminoglycan linkages.

Findings from our laboratory and others have shown that IAIP are important components of the innate immune response that modulate host responses to pathological insults^10–11,19,21^. It is known that IAIP, as part of the innate immune response, protect against the damaging effects of proteases released during acute systemic inflammation following severe infections, burns, trauma and injury. As a consequence, these proteins are rapidly consumed and excreted, leading to a rapid decrease in plasma levels. We previously reported that circulating IAIP levels are significantly decreased in adult and newborn sepsis and that the total IAIP levels correlate inversely with the mortality rate in patients with severe sepsis^12,22,29^. Similar reductions have been observed in liver-derived plasma proteins, such as apolipoprotein A-I (apoA-I), histidine-rich glycoprotein (HRG), kininogen-1 (KNG-1), and vitronectin (VN) in patients with sepsis due to systemic inflammation^30^. It is known that the hepatic biosynthesis of IAIP, declines with systemic immune system activation and as such IAIP are widely considered an acute phase reactant^31^. Importantly, in several adult and newborn animal models of sepsis and anthrax infection, IAIP replacement therapy has been demonstrated to overcome the depletion of IAIP levels and reduce sepsis related mortality^32–34^.

Our study validates IAIP as a reliable biomarker for these conditions, with LFIA providing a simple, rapid, accurate, and clinically practical alternative to ELISA. IAIP measured by LFIA demonstrated an 80.0% sensitivity and 92.3% specificity for the detection of neonatal sepsis; IAIP LFIA was 84.6% sensitive and 86.7% specific for the detection of NEC. These results, and the very high positive and negative predictive values, suggest that measurement of IAIP may play an important role in clinical decisions regarding antibiotic therapy. These results combined with our previous publications^12,25,29^ which include almost 900 infants suggest that reference ranges of IAIP can be defined (Table 3). Normal circulating IAIP is greater than 300 µg/ml. Levels that are mildly, moderately and severely reduced are seen at 200-300, 100-200, <100 ug/ml respectively.

**Table 3.** Reference ranges of IAIP (the lower limit, µg/ml) in infants.

|  |  |
| --- | --- |
| IAIP ≥ 300 | Normal |
| 200-300 | Mildly decreased |
| 100-200 | Moderately decreased |
| < 100 | Severely decreased |

**Table 4.** Summary of the indicators of performance of IAIP by ELISA and LFIA.

|  | Threshold<br>(µg/ml) | AUC | Sensitivity<br>(%) | Specificity<br>(%) | Accuracy<br>(%) | PPV<br>(%) | NPV<br>(%) |
| --- | --- | --- | --- | --- | --- | --- | --- |
| Sepsis ELISA | 126 | 0.893 | 94.2 | 71.4 | 85.1 | 83.1 | 89.3 |
| Sepsis LFIA | 118 | 0.925 | 80.0 | 92.3 | 87.4 | 87.5 | 87.3 |
| NEC ELISA | 124 | 0.872 | 93.3 | 69.2 | 82.1 | 77.8 | 90.0 |
| NEC LFIA | 144 | 0.887 | 84.6 | 86.7 | 85.7 | 84.6 | 86.7 |

AUC: area under the curve; PPV: positive predictive value; NPV: negative predictive value

Fifty-six percent of very low birth weight infants receive empiric antibiotics for treatment^35^. However, the deleterious effects of prolonged antibiotics exposure are widely appreciated^36^. Prolonged use of empiric antibiotics is associated with an increased risk of NEC and death in extremely low birth weight infants^37–38^. Propensity score testing demonstrates that critically ill infants receiving very broad-spectrum empiric antibiotic coverage for respiratory failure have worse outcomes than infants receiving narrower coverage^39^. They have a higher incidence of late-onset sepsis which is often associated with significant antibiotic resistance^39^. As a biomarker for early and accurate identification of sepsis and/or NEC, IAIP may be very useful in the challenging decisions on initial use, continuation or early termination of antibiotic treatment in infants. Our case controls were identified with a clinical presentation consistent with sepsis or NEC. However subsequent clinical course showed rapid resolution allowing discontinuation of antibiotics and resumption of feeds. The very high, demonstrated reliability of the LFIA, when compared to the reference ELISA, suggests that, with rapid results of IAIP measurement, even stricter criteria for antibiotic administration may be feasible in this clinical setting. Additional studies are warranted.

A new observation in this report is that infants with “clinical sepsis” (commonly referred to as “suspected sepsis”, “culture-negative sepsis”, “presumed sepsis”) have comparable reductions in circulating IAIP levels to infants with confirmed sepsis and NEC. In adults one nominal blood sample up to 20 ml is used to inoculate two bottles and a standard set of two blood cultures are necessary to identify a pathogen in 80% to 96% of bacteremias^40–42^. For infants, it is difficult to collect comparable blood samples. As a result, neonatologists are faced with a greater problem of false-negative blood cultures. The terms “suspected sepsis”, “culture-negative sepsis”, “presumed sepsis” and “clinical sepsis” are widely used in the NICU to describe sick infants without culture-proven evidence of a pathogen. In 2016, the National Institute for Health and Care Excellence (NICE) published guidance on recognizing, diagnosing, and managing suspected sepsis. The report suggested using an inflammatory biomarker like C-reactive protein (CRP) in assistance to diagnose suspected sepsis^43^. Our study shows that reduced circulating IAIP levels are a feature of the infants with “clinical sepsis” and IAIP levels are a potentially reliable biomarker in this setting. Moreover, we have shown that IAIP level is more reliable than CRP in the diagnosis of sepsis, SIP and NEC^25,44^.

## Limitations

We collected our samples at the time of an acute event and then any additionally available residual samples over 14 days. This was a prospective observational study but we did not have IRB and parental consent for systematic collection of blood samples following clinical presentation. The decision for blood collection, and thus availability of residual blood, was entirely up to the clinical teams who ordered more tests in the first few subsequent days. As a result, there were not enough samples for robust comparison of levels that were obtained more than 72 hours after acute events. Another limitation of this work is that the study period overlapped with COVID-19 pandemic. Recruiting subjects was difficult and thus the sample size was relatively modest.

## Conclusions

This study highlights IAIP as a promising biomarker for neonatal sepsis and NEC, with LFIA offering a simple, rapid, accurate, and clinically viable alternative to ELISA. Given the high sensitivity and specificity of IAIP measurement, its integration into clinical workflows may help improve early diagnosis, optimize antibiotic stewardship, and reduce unnecessary feeding interruptions. Further research is warranted to validate these findings in larger cohorts.

## Data Availability

Data will be available upon request to the corresponding author.

## Acknowledgements

We thank our core labs for their hard work, Ross S Price, Director of IRB Administration, Women & Infants Hospital of Rhode Island, for providing feedback; Erin Bohon, LPN, Clinical Research Program Coordinator and Jeffrey Eckert, Ph.D., Center for Pregnancy and Newborn Health at the University of Oklahoma Health Sciences, for recruitment, collection, storage and shipment of specimens.

